# Impact of Rotavirus Vaccine on Malnutrition Among Children in India—Breaking Vicious Cycle of Diarrhea: A Cross-Sectional Analytical Study

**DOI:** 10.64898/2026.02.05.26345714

**Authors:** Ajay Kumar Verma, Pritu Dhalaria, Pawan Kumar, Sanjay Kapur, Pretty Priyadarshini, Ajeet Kumar Singh, Kapil Singh, Bhupendra Tripathi, Arindam Ray

**Affiliations:** Immunization Technical Support Unit, Ministry of Health & Family Welfare, Government of India, New Delhi,110070, India; Immunization Division, Ministry of Health & Family Welfare, New Delhi 110011, India; John Snow India, New Delhi, 110070, India; Gates Foundation, New Delhi, 110067, India

**Keywords:** Rotavirus, Vaccine, Malnutrition, Diarrhea, Stunting, Wasting, Impact

## Abstract

**Background:** Malnutrition among children remains a significant public health concern in countries with stunting, wasting, and underweight as key indicators. Rotavirus accounts for approximately 40% of moderate to severe diarrhea cases in children under five, highlighting the potential role of rotavirus vaccination in mitigating diarrhea-associated malnutrition. This study investigated the association between the Rotavirus vaccine (RVV) and malnutrition among children in India.

**Methods:** The study examined data from the National Family Health Survey-5. The sample included 67,369 children aged 12 to 35 months. Outcomes included stunting, underweight, and wasting, along with their severe form of malnutrition. Adjusted regression, sensitivity analyses, and Inverse Probability Weighting Regression Adjustment assessed the association between receiving RVV and malnutrition.

**Results:** Children receiving all three RVV doses had a lower prevalence of stunting (37% vs 41%), underweight (28% vs 33%), and wasting (17% vs 20%) compared to the unvaccinated. Full RVV coverage was significantly associated with reduced odds of stunting (aOR: 0.88; 95% CI: 0.85–0.92), underweight (aOR: 0.86; 95% CI: 0.83–0.89), and wasting (aOR: 0.85; 95% CI: 0.82–0.89).

**Conclusion:** Receiving RVV is associated with a reduced risk of malnutrition in children, highlighting the indirect effect (Herd Immunity) and the role of the vaccine beyond preventing diarrheal disease.

## 1. Introduction

Malnutrition is a widely recognized public health challenge that affects all regions worldwide. It refers to imbalances in a person’s nutritional intake, both deficiencies and excesses—undernutrition and overnutrition. Within these two broad categories, undernutrition comprises stunting, wasting, and underweight, the three key indicators of malnutrition. Malnutrition in children is a critical indicator that measures a child’s nutritional status and health. Among children under five years of age (CU5), malnutrition is driven by the intricate interaction of factors, including the availability of age-appropriate, nutrient-rich, and sufficient food, adherence to hygiene practices, accessibility and utilization of healthcare services, and living environments that enable healthy dietary behaviours[1]. There has been considerable progress in recent decades. The worldwide prevalence of stunting in CU5 declined from 33% (204.2 million) in 2000 to 22.3% (148.1 million) in 2022. Similarly, the wasting rate decreased from 8.7% to 6.8% (45 million), and undernutrition decreased from 20.5% to 12.6% between 2020 and 2022[2].

Malnourished children have multiple immediate and long-term adverse consequences, including a weakened immune system and bodily function, low cognitive and motor development, and increased vulnerability to diseases. Children with multiple anthropometric failures, particularly those who are stunted, underweight, and wasted, are linked with higher mortality risks[3]. Malnutrition is a major contributor to child mortality in low– and middle-income countries (LMICs), accounting for nearly half of the deaths among CU5, often in combination with infectious diseases[4–7]. Malnutrition is not limited to changes in dietary phenomena and is rather complex. Child malnutrition and diarrhea have a bidirectional relationship. Malnutrition is both a cause and a consequence of diseases like diarrhea[8,9]. Malnutrition leaves children more susceptible to infections and diarrheal episodes, and diarrhea further alters their nutritional state, leaving them vulnerable to recurrent infections (Figure 1). It is a vicious cycle of infectious diseases and malnutrition[10]. The key indicators stunting, wasting, and underweight increase with the severity and frequency of infections and diarrheal episodes. Stunting results from long-term nutritional deprivation and recurrent and chronic infections, resulting in diminished human capital and economic potential compared to children with normal stature[11,12]. Evidence suggests that frequent diarrheal diseases in early childhood impact a child’s mental development later in their lives, affecting school readiness and performance, either directly or indirectly as a result of stunting. It also increases susceptibility to infections and non-communicable diseases later in life[13,14].

**Figure 1:**
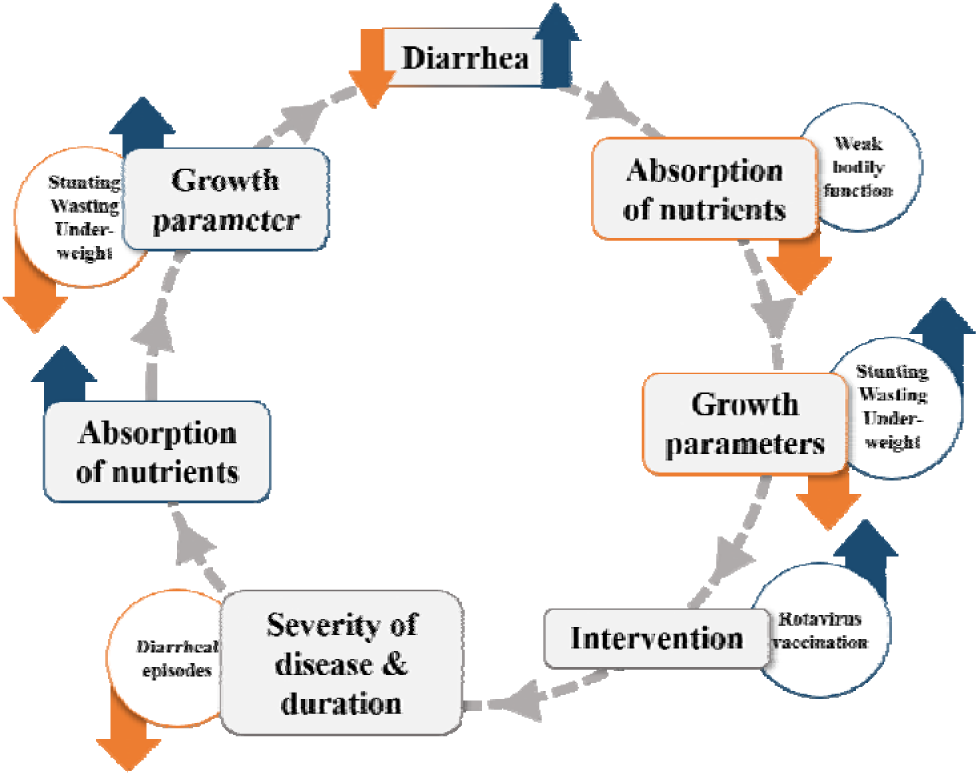
Bi-directional relationship and synergistic cycle of diarrhea and malnutrition. In this figure, the blue and orange arrows indicate an increase and a decrease, respectively. With the onset of diarrhea in children, nutritional absorption decreases, affecting growth and increasing stunting, wasting, and underweight. Rotavirus vaccines (RVV) play a critical role in breaking the vicious cycle between diarrhea and malnutrition. As RVV coverage increases, the severity, number of episodes, and cases of diarrhea reduce, leading to improved nutrition absorption. As a result, there is a drop in stunting, wasting, and underweight cases, leading to a reduction in diarrhea prevalence. This illustrates the bi-directional relationship and the vicious/synergistic cycle between the two.

Diarrheal disease predominates over all other non-dietary causes of malnutrition in low-income countries, reducing dietary intake, leading to malabsorption, and poor digestion[9,15]. Malabsorption generally plays a central role in the interaction between malnutrition and chronic diarrhea, resulting in both nutritional deficits and diarrhea. With severe malnutrition, chronic diarrhea can persist due to impaired immune function and poor mucosal recovery. The combination and vicious cycle of malnutrition and diarrhea becomes lethal due to their interaction with each other. A 2013 estimation highlights that every year in India, rotavirus is responsible for nearly 40% of moderate to severe diarrhea in children under five, amounting to 872,000 hospitalizations, 3,270,000 outpatient visits, and 78,000 deaths[16,17]. Rotavirus has the largest attributable fraction (AF) among pathogens causing diarrhea in children aged 12-23 months, despite a decrease in AF with age[18].

Undernutrition and infectious diseases are interrelated global health challenges, particularly affecting children in developing countries[19,20]. Several interventions have been implemented in the last decade to reduce malnutrition and childhood diarrhea. The World Health Organization (WHO) advocates integrating rotavirus vaccination into national immunization programs, particularly in regions with a high incidence of rotavirus-induced diarrhea, such as Asia and Sub-Saharan Africa. In line with this recommendation, in 2016 India introduced an indigenous monovalent rotavirus vaccine (RVV) into the Universal Immunization Program (UIP), making it the first country in the South-East Asia Region (SEAR) to launch this vaccine. The RVV began as a pilot in select states and was gradually expanded in phases, achieving nationwide rollout by 2019. India’s National Family Health Survey-5 (NFHS) data from 2019-21 indicated that approximately 7% of children under 5 years had episodes of diarrhea two weeks prior to the survey. The prevalence of diarrhea has decreased from 9% to 7% between NFHS-4 (2015-16) and NFHS-5 (2019-21). The Prime Minister’s Overarching Scheme for Holistic Nutrition (POSHAN) Abhiyaan, known as the National Nutrition Mission, is a flagship program of the government that aims to improve the nutritional status of children, pregnant women, and lactating mothers. Launched in 2018, the mission directs the country’s attention to malnutrition. An integral strategy of this mission is to expand the RVV coverage nationwide and reduce the prevalence of Rotavirus-induced diarrhea[21].

Evidence shows that children who received the RVV had a lower risk of diarrhea than those who did not[17,22]. Rotavirus vaccination has significantly improved child health by reducing diarrheal illnesses and their associated complications, resulting in a substantial reduction in mortality among children under five years globally. The vaccine has prevented 139,000 under-five deaths between 2006 and 2019. In 2019 alone, the vaccine prevented 15% of under-five deaths[23]. In LMICs, rotavirus vaccination has been associated with a more than 40% reduction in all-cause diarrheal mortality in children[24–26]. Growing evidence indicates that diarrhea significantly impacts childhood growth and cognitive development, with a strong association between diarrheal diseases and undernutrition[27–30]. RVV protects children against various diarrheal diseases, thereby improving their physical growth and development[31,32]. The Expanded Program on Immunization (EPI) has been proven highly effective in reducing child mortality, thereby increasing life expectancy by preventing infectious diseases across various settings[33,34]. Emerging evidence indicates that the impact of vaccines extends beyond the prevention of target pathogens to other, unrelated pathogens known as *’heterologous effects’*. RVV has beneficial effects on other pathogens, likely through its immune-stimulatory effects[35–37]. In addition, immunized individuals receive direct protection against specific pathogens, while high vaccination rates provide indirect protection, known as herd immunity, to unvaccinated individuals in the population[38,39]. By reducing the frequency of disease in undernourished children, RVV helps maintain their nutritional status and use existing calories for growth and cognitive development.

Malnutrition among children under five continues to receive policy attention in India. According to NFHS-5 (2019-21), the prevalence of stunting is 36%, wasting is 19%, and overall underweight stands at 32%. These figures represent a gradual improvement from NFHS-4 (2015-16), where stunting was 38.4%, wasting 21.0%, and underweight 35.7%, highlighting the progress made through sustained national programs[40]. To date, evidence has been lacking on the effect of RVV introduction on diarrhea-mediated growth faltering. The indirect impact of the RVV on reducing childhood malnutrition has not received adequate attention in scientific discourse or policy decision-making. While the vaccine’s primary role in lowering under-five mortality and morbidity from diarrheal diseases is well established, its critical secondary benefit, contributing to improved nutritional status, remains underrecognized. Our hypothesis states that rotavirus vaccination reduces the risk of rotavirus-induced diarrhea directly and indirectly through other pathogen-associated diarrhea, and nutrient malabsorption, thereby contributing to improved anthropometric outcomes, such as height-for-age, weight-for-height, and weight-for-age metrics, in vaccinated children compared to those who are unvaccinated. This study aims to investigate the association between rotavirus vaccination and malnutrition indicators (stunting, wasting, and underweight) among children aged 12 to 35 months in India, using nationally representative NFHS-5 data. The research aligns with the country’s policy priorities, including the National Nutrition Mission, which emphasizes nutrition and immunization as key strategies to reduce child malnutrition. This mission has a dual imperative: to prevent rotavirus infections and decisively reduce malnutrition associated with diarrheal incidences. By examining how rotavirus vaccination may improve nutritional outcomes by preventing diarrheal disease, this study aims to generate evidence that bridges immunization and nutrition policy efforts, adding a novel perspective to the scientific literature.

## 2. Data and methods

### 2.1 Data source

The National Family Health Survey 2019-21 (NFHS-5) is the fifth in the NFHS series, following the first conducted in 1992-93. NFHS provides comprehensive data on health indicators and socioeconomic characteristics of women, children, and men, enabling policymakers to evaluate the progress and status of health conditions over time. NFHS-5 specifically focused on key areas, including infant and child mortality, maternal and child health, fertility, and other important health and welfare indicators in India at the national, state, and district levels. It employed a national, state/union territory (UT), and district-representative sample survey. The survey used a stratified two-stage sampling design to collect data from eligible populations and respondents. Data were collected on health indicators from 724,115 women and 101,839 men across 636,699 households and 30,198 primary sampling units. NFHS-5 was conducted in two phases: Phase I (17 June 2019 to 30 January 2020), covering 17 states and 5 UTs, and Phase II (2 January 2020 to 30 April 2021), encompassing 11 states and 3 UTs. NFHS-5 also provides information on the health status and health services of children <5 years (subpopulation), among whom 724,115 women aged 15-49 years gave birth in the five years preceding the survey. A total of 67,979 children aged 12 to 35 months were included in the analysis, following adjustment for relevant background variables.

### 2.2 Variables

The outcome variables included wasting, stunting, and underweight among children aged 12-35 months, following the WHO 2019 guidelines[41]. Stunting is defined as the percentage of children under the age of five whose height-for-age Z-score (HAZ) is below –2 and –3 standard deviations (SD) from the median height-for-age of a reference population (Height-for-age < –2SD and < –3SD). Underweight refers to children whose weight-for-age Z-score (WAZ) is below –2 and –3 SD from the median weight-for-age of the reference population (Weight-for-age < –2SD and < –3SD). Wasting is characterized by the percentage of children whose weight-for-height Z-score is below –2 and –3 SD from the median weight-for-height of the reference population (Weight-for-height < –2SD and < – 3SD). Furthermore, we generated three additional outcome variables related to concurrently present diarrhea and malnutrition, which include diarrhea and Height-for-age < –2SD, diarrhea and Weight-for-age < –2SD, and diarrhea and Weight-for-height < –2SD, where information about diarrhea in children was collected over the two weeks before the survey.

The NFHS-5, conducted from 2019 to 2021, provides, for the first time, data on rotavirus vaccination for children under five years in India. This data is a key explanatory variable in this analysis. Information about rotavirus vaccination was gathered from vaccination cards and maternal reports/recalls. As part of India’s UIP, three doses of the RVV are administered at 6 weeks, 10 weeks, and 14 weeks, and the complete vaccination schedule should be completed by one year of age. In addition to rotavirus vaccination status, other explanatory variables included in the analysis are the child’s age, sex, birth order, maternal education level, maternal body mass index (BMI), religion, place of residence, social group (caste), place of delivery (institutional or home), wealth quintile, and type of toilet facilities. These are key sociodemographic variables that impact the health indicators.

### 2.3 Empirical Strategy

In this study, we conducted a simple cross-tabulation analysis of height-for-age, weight-for-age, and weight-for-height in relation to the RVV administration among children aged 12-35 months. This study used univariate and multivariate logistic regression analyses to evaluate the influence of the rotavirus vaccination on undernutrition outcomes in children. Specifically, we focused on anthropometric indicators including height-for-age below –2 standard deviations (SD) and –3 SD, weight-for-age below –2 SD and –3 SD, and weight-for-height below –2 SD and –3 SD. The analysis employed six multivariate logistic regressions on anthropometric measures and rotavirus vaccines. The model is specified as follows.

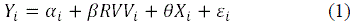

In this context, *Y_i_* represents the undernutrition outcomes for child *i*, which include stunting, wasting, or underweight. The term Cl*_i_* denotes the intercept, while *RVV_i_* signifies the primary explanatory variable, indicating whether children received one or more doses of the RVV between the ages of 12-35 months. The variable *X_i_* encompasses a variety of control variables, such as the child’s age, gender, household socioeconomic status, maternal education, and other relevant factors. The □*_i_* term denotes error.

We employed three additional multivariate logistic regressions to examine the impact of the RVV on the concurrent burden of diarrheal infection and undernutrition (children who experience both diarrhea and stunting, wasting, and underweight). In equation (1), replace the outcome *Y_i_* with the concurrent incidence of diarrhea and malnutrition indicators to represent those children who experience diarrhea and undernutrition simultaneously for child *i*.

The objective was to understand how the RVV influences undernutrition among children. Mediation involves considering a mediator that explains how an independent variable impacts an outcome. This is crucial in treatment studies, as it helps us understand how an intervention achieves its desired effect[42]. Mediation analysis examines how an independent variable influences a dependent variable through an intervening variable, providing deeper insights into these relationships[43]. Our study examined the indirect effects of rotavirus vaccination on childhood undernutrition to identify potential pathways involved. We focused on diarrhea as a significant mediator, due to its established role in disrupting nutrient absorption and contributing to growth failure (Figure 2). Diarrhea was selected for analysis because it is likely to mediate the hypothesized causal mechanism between rotavirus vaccination and undernutrition outcomes (stunting, underweight, and wasting).

**Figure 2:**
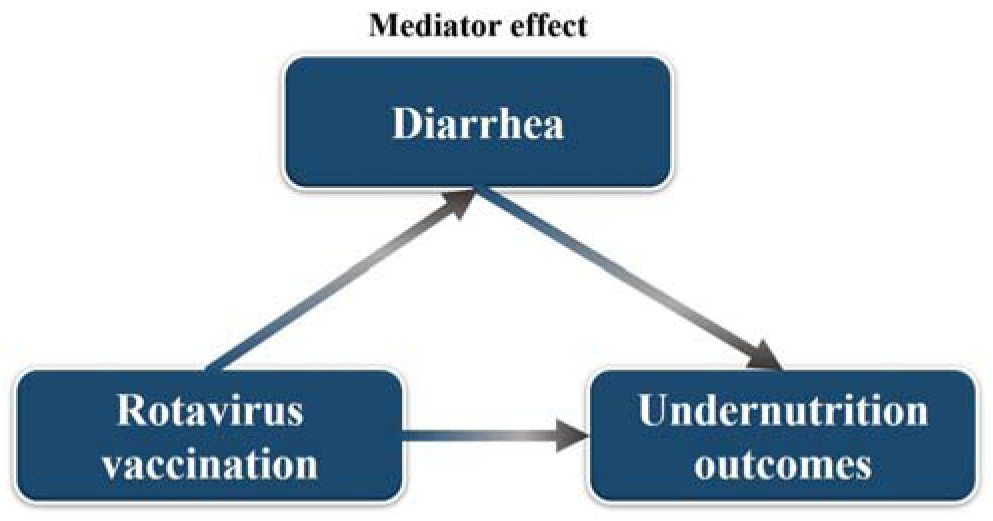
Interaction between Rotavirus vaccination, diarrhea, and undernutrition outcomes.

We employed the Generalized Structural Equation Model (GSEM) to estimate the indirect relationship between RVV, diarrhea, and the undernutrition status of children aged 12 to 35 months. There are advantages to using the GSEM approach in mediation analysis, particularly when the model contains categorical, binary, ordered, count, etc., variables, which allow easy interpretation and estimation[44]. First, we used the two-level model with the GSEM command and the logistic regression approach in Stata to analyse the structural pathways, followed by the nlcom command to estimate the indirect effects of rotavirus vaccination on childhood undernutrition.

In addition to evaluating the more reliable, bias-free impact of the rotavirus vaccine on malnutrition outcomes in children using observational data, we used the Inverse Probability Weighted Regression Adjustment (IPWRA) method. This method involves weighting each observation by its propensity score. By combining model regression adjustment (RA) and inverse probability weighting (IPW), IPWRA estimates the average treatment effect on the treated (ATT) and the average treatment effect (ATE) from observational data. IPWRA estimators use weighted regression coefficients to compute averages of treatment-level predicted outcomes, where the weights are the estimated inverse probabilities of treatment. The differences between these averages provide estimates of the treatment effects. IPWRA estimators are known for their double-robust property[45]. We used two models to assess the treatment effect of the rotavirus vaccine on malnutrition outcomes. In the first model, children who received treatment are defined as those who received either one or two doses of the rotavirus vaccine out of three, while untreated children didn’t receive any dose. In the second model, children who received treatment are defined as those who completed all three doses of the rotavirus vaccine, and untreated children didn’t receive any dose.

## 3. Results

### 3.1 Diarrhea and Malnutrition

Figure 3 illustrates that children aged 12 to 35 months who received the rotavirus vaccine had a lower prevalence of stunting, wasting, and underweight than those who did not. This remains true to malnutrition (below –2SD) and its severe form (below –3SD).

**Figure 3:**
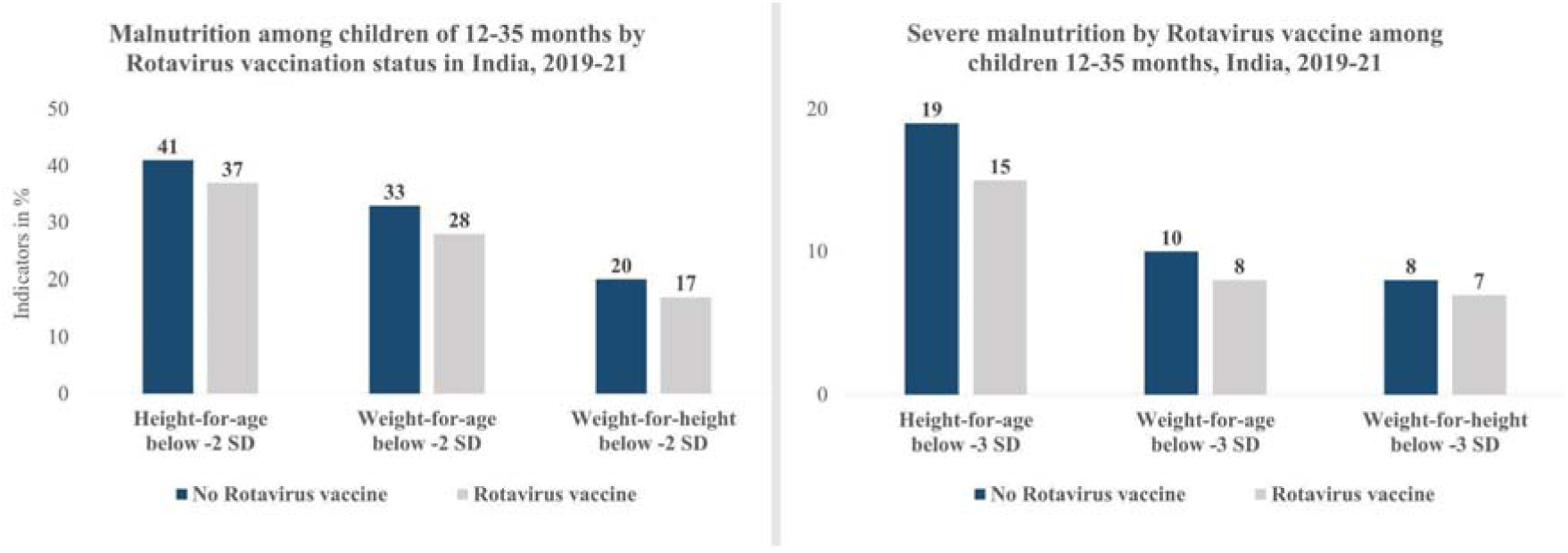
Malnutrition (below –2 SD) and severe malnutrition (below –3 SD) among children of 12-35 months by Rotavirus vaccination status in India, 2019-21.

### 3.2 Sample Characteristics and Sample Distribution

Table 1: Among children aged 12-35 months in our study, 8.3% reported having diarrhea in the two weeks prior to the survey. Regarding vaccination, 59% received no doses, 6.6% received 1 or 2 doses, and 34.4% received all 3 doses of the rotavirus vaccine. Also, 38.9% were stunted, 30.6% were underweight, and 19.1% were wasted.

**Table 1:**
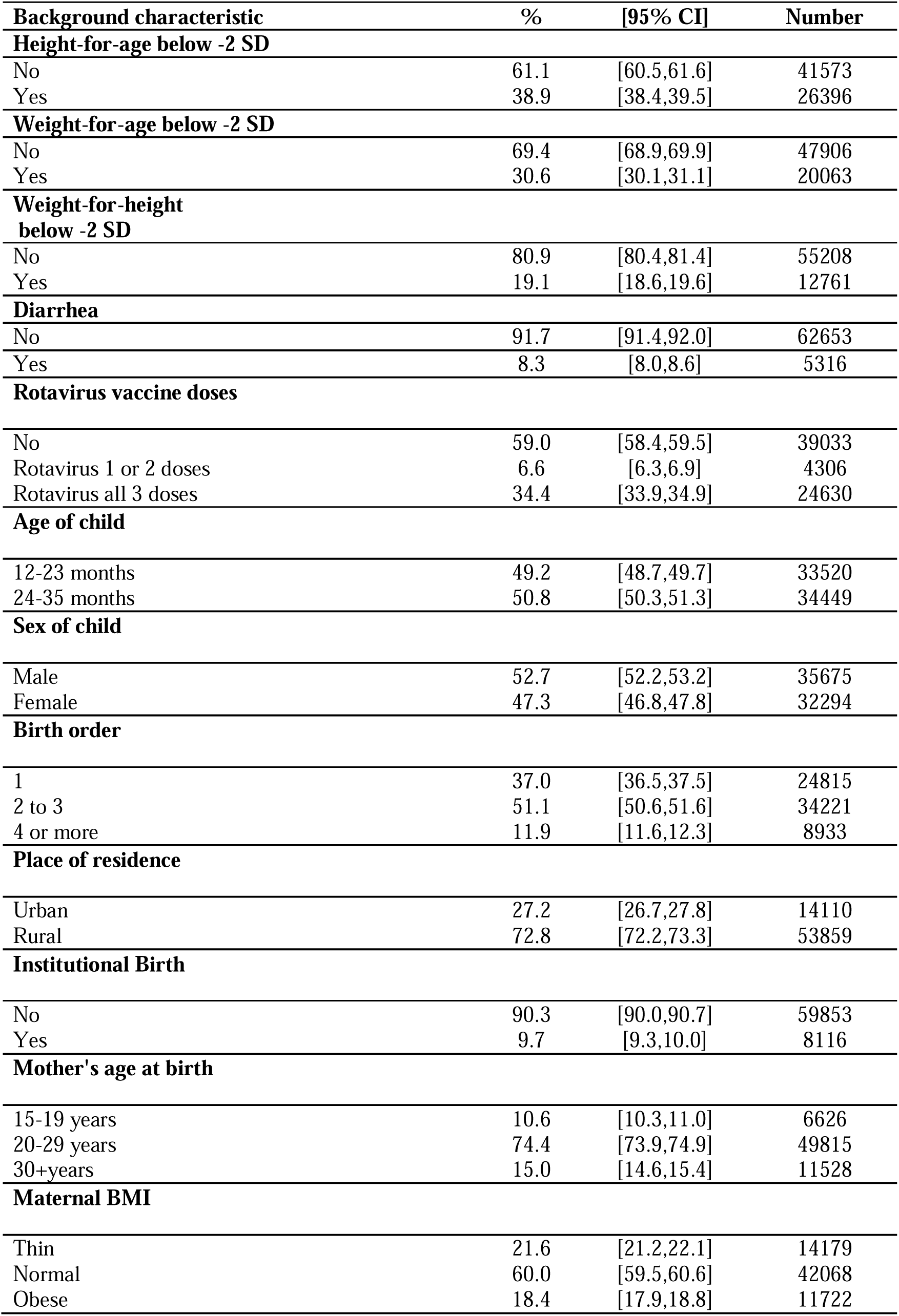
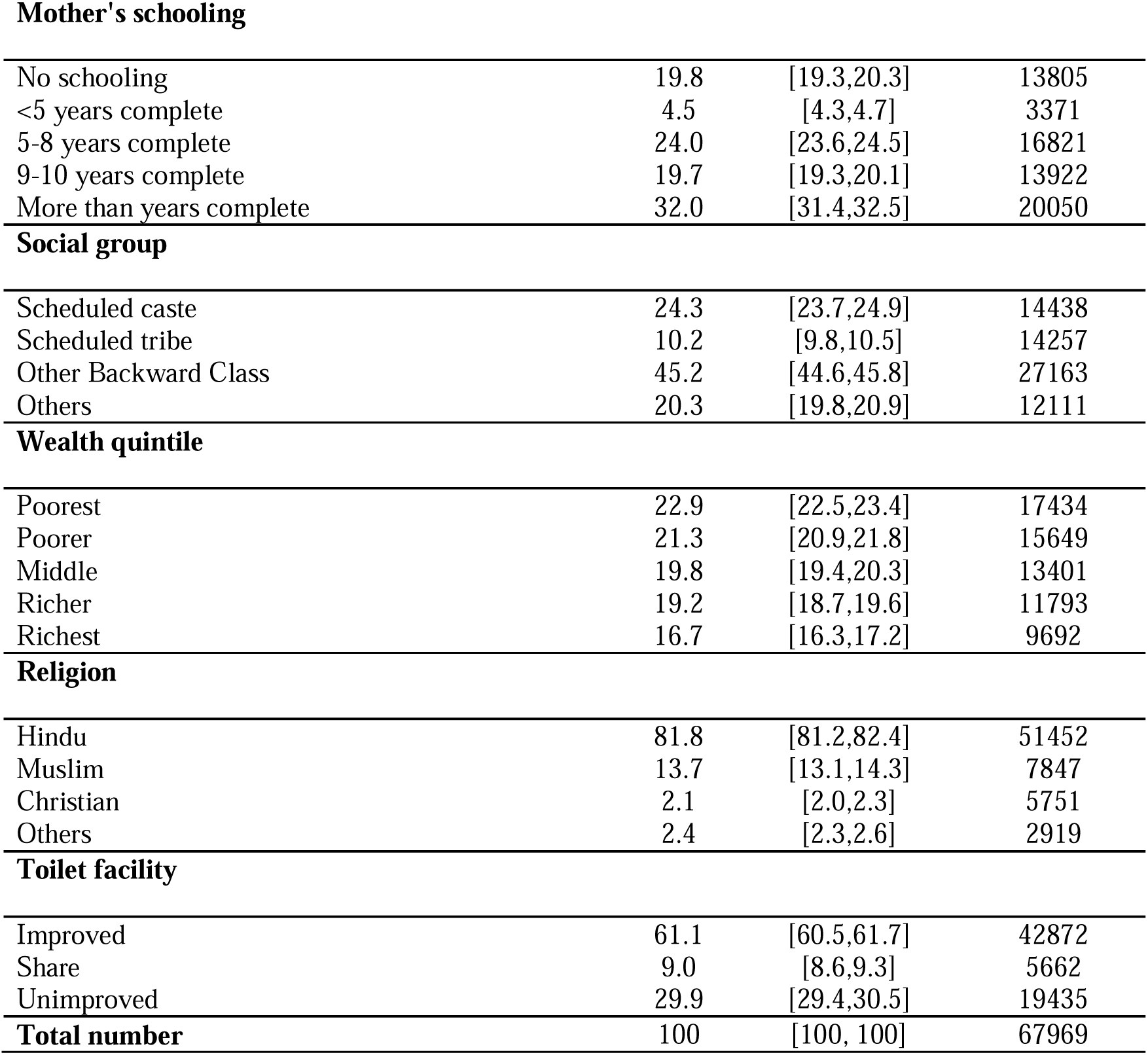
Sample Distribution of the variables among 12-35 months old children, India, 2019-21.

| **Background characteristic** | **%** | **[95% CI]** | **Number** |
| --- | --- | --- | --- |
| **Height-for-age below -2 SD** |  |  |  |
| No | 61.1 | [60.5,61.6] | 41573 |
| Yes | 38.9 | [38.4,39.5] | 26396 |
| **Weight-for-age below -2 SD** |  |  |  |
| No | 69.4 | [68.9,69.9] | 47906 |
| Yes | 30.6 | [30.1,31.1] | 20063 |
| **Weight-for-height below -2 SD** |  |  |  |
| No | 80.9 | [80.4,81.4] | 55208 |
| Yes | 19.1 | [18.6,19.6] | 12761 |
| **Diarrhea** |  |  |  |
| No | 91.7 | [91.4,92.0] | 62653 |
| Yes | 8.3 | [8.0,8.6] | 5316 |
| **Rotavirus vaccine doses** |  |  |  |
| No | 59.0 | [58.4,59.5] | 39033 |
| Rotavirus 1 or 2 doses | 6.6 | [6.3,6.9] | 4306 |
| Rotavirus all 3 doses | 34.4 | [33.9,34.9] | 24630 |
| **Age of child** |  |  |  |
| 12-23 months | 49.2 | [48.7,49.7] | 33520 |
| 24-35 months | 50.8 | [50.3,51.3] | 34449 |
| **Sex of child** |  |  |  |
| Male | 52.7 | [52.2,53.2] | 35675 |
| Female | 47.3 | [46.8,47.8] | 32294 |
| **Birth order** |  |  |  |
| 1 | 37.0 | [36.5,37.5] | 24815 |
| 2 to 3 | 51.1 | [50.6,51.6] | 34221 |
| 4 or more | 11.9 | [11.6,12.3] | 8933 |
| **Place of residence** |  |  |  |
| Urban | 27.2 | [26.7,27.8] | 14110 |
| Rural | 72.8 | [72.2,73.3] | 53859 |
| **Institutional Birth** |  |  |  |
| No | 90.3 | [90.0,90.7] | 59853 |
| Yes | 9.7 | [9.3,10.0] | 8116 |
| **Mother's age at birth** |  |  |  |
| 15-19 years | 10.6 | [10.3,11.0] | 6626 |
| 20-29 years | 74.4 | [73.9,74.9] | 49815 |
| 30+years | 15.0 | [14.6,15.4] | 11528 |
| **Maternal BMI** |  |  |  |
| Thin | 21.6 | [21.2,22.1] | 14179 |
| Normal | 60.0 | [59.5,60.6] | 42068 |
| Obese | 18.4 | [17.9,18.8] | 11722 |
| **Mother's schooling** |  |  |  |
| No schooling | 19.8 | [19.3,20.3] | 13805 |
| <5 years complete | 4.5 | [4.3,4.7] | 3371 |
| 5-8 years complete | 24.0 | [23.6,24.5] | 16821 |
| 9-10 years complete | 19.7 | [19.3,20.1] | 13922 |
| More than years complete | 32.0 | [31.4,32.5] | 20050 |
| **Social group** |  |  |  |
| Scheduled caste | 24.3 | [23.7,24.9] | 14438 |
| Scheduled tribe | 10.2 | [9.8,10.5] | 14257 |
| Other Backward Class | 45.2 | [44.6,45.8] | 27163 |
| Others | 20.3 | [19.8,20.9] | 12111 |
| **Wealth quintile** |  |  |  |
| Poorest | 22.9 | [22.5,23.4] | 17434 |
| Poorer | 21.3 | [20.9,21.8] | 15649 |
| Middle | 19.8 | [19.4,20.3] | 13401 |
| Richer | 19.2 | [18.7,19.6] | 11793 |
| Richest | 16.7 | [16.3,17.2] | 9692 |
| **Religion** |  |  |  |
| Hindu | 81.8 | [81.2,82.4] | 51452 |
| Muslim | 13.7 | [13.1,14.3] | 7847 |
| Christian | 2.1 | [2.0,2.3] | 5751 |
| Others | 2.4 | [2.3,2.6] | 2919 |
| **Toilet facility** |  |  |  |
| Improved | 61.1 | [60.5,61.7] | 42872 |
| Share | 9.0 | [8.6,9.3] | 5662 |
| Unimproved | 29.9 | [29.4,30.5] | 19435 |
| **Total number** | 100 | [100, 100] | 67969 |

### 3.3 Regression Analysis of malnutrition and rotavirus vaccination

Table 2 provides adjusted odds ratios (aORs) and 95% confidence intervals (CIs) for the association between various background characteristics and the likelihood of children having height-for-age, weight-for-age, and weight-for-height below –2 standard deviations (SD). The results indicate that children who received one or two doses of the RVV had lower odds of being stunted (aOR: 0.94, 95% CI: 0.88–1.00), being underweight (adjusted odds ratio (aOR: 0.91, 95% CI: 0.85–0.98), and experiencing wasting (aOR: 0.89, 95% CI: 0.82–0.96). Additionally, those who received three doses showed significantly reduced odds of stunting (aOR: 0.88, 95% CI: 0.85–0.92), underweight (aOR: 0.86, 95% CI: 0.83–0.89), and wasting (aOR: 0.85, 95% CI: 0.81–0.88) compared with those who did not receive any dose. Girls had lower odds of stunting (aOR: 0.86, 95% CI: 0.84–0.89), underweight (aOR: 0.88, 95% CI: 0.85–0.91), and wasting (aOR: 0.90, 95% CI: 0.86–0.93) compared with boys. Children aged 24-35 months had higher odds of being underweight (aOR: 1.17, 95% CI: 1.13–1.21) but slightly lower odds of stunting (aOR: 0.96, 95% CI: 0.93–0.99) compared with children aged 12-23 months. Birth order was a significant risk factor, with children in fourth or higher birth order showing increased odds of stunting (aOR: 1.47, 95% CI: 1.38–1.56) and underweight (aOR: 1.40, 95% CI: 1.31–1.50) compared with first order children. Children in rural areas were less likely to experience stunting (aOR: 0.95, 95% CI: 0.91–1.00), underweight (aOR: 0.89, 95% CI: 0.85–0.94), and wasting (aOR: 0.84, 95% CI: 0.80–0.88) compared to those in urban areas. Non-institutional births were associated with higher odds of stunting and underweight (aOR: 1.11 for both, 95% CI: 1.05–1.18). Children born to mothers aged 30 years or older had significantly lower odds of stunting (aOR: 0.79, 95% CI: 0.73–0.85) and underweight (aOR: 0.90, 95% CI: 0.84–0.97) but higher odds of wasting (aOR: 1.17, 95% CI: 1.07–1.27) compared to mothers aged 15–19 years. Maternal BMI emerged as a significant protective factor, as children of mothers with normal BMI (18.5-24.9 kg/m2) had reduced odds of stunting (aOR: 0.73, 95% CI: 0.70–0.76), underweight (aOR: 0.61, 95% CI: 0.58–0.63), and wasting (aOR: 0.79, 95% CI: 0.76–0.83). Maternal education also played a protective role, with mothers who completed more than 10 years of schooling showed significantly reduced odds of stunting (aOR: 0.67, 95% CI: 0.63–0.70), underweight (aOR: 0.72, 95% CI: 0.68–0.77), and wasting (aOR: 0.86, 95% CI: 0.81–0.93) compared to those with no schooling. Children from the richest quintile had the lowest odds of stunting (aOR: 0.54, 95% CI: 0.50–0.58), underweight (aOR: 0.48, 95% CI: 0.44–0.52), and wasting (aOR: 0.73, 95% CI: 0.67–0.80) compared to the poorest quintile. Children from scheduled tribes had higher odds of wasting (aOR: 1.21, 95% CI: 1.13–1.30), while Muslim children were more likely to experience stunting (aOR: 1.11, 95% CI: 1.05–1.16) and wasting (aOR: 1.12, 95% CI: 1.05–1.18). Use of unimproved toilet facilities was associated with higher odds of stunting and underweight (aOR: 1.09, 95% CI: 1.05–1.14 for both) and wasting (aOR: 1.04, 95% CI: 0.99–1.09). Additionally, results for the association between various background characteristics and the likelihood of anthropometric measurements below –3 standard deviations (SD), with adjusted odds ratios (aORs), are reflected in Supplementary Table 1. Supplementary Table 2 demonstrates a positive association between rotavirus vaccination and anthropometric measurements below –2 standard deviations (SD) compared with those who did not receive any doses among children aged 12-23 months who had received all basic vaccinations. Supplementary Table 3 shows multivariate logistic regression results for Rotavirus vaccine and undernutrition among children below –2 SD and whose vaccine was reported by the card by sociodemographic characteristics.

**Table 2:**
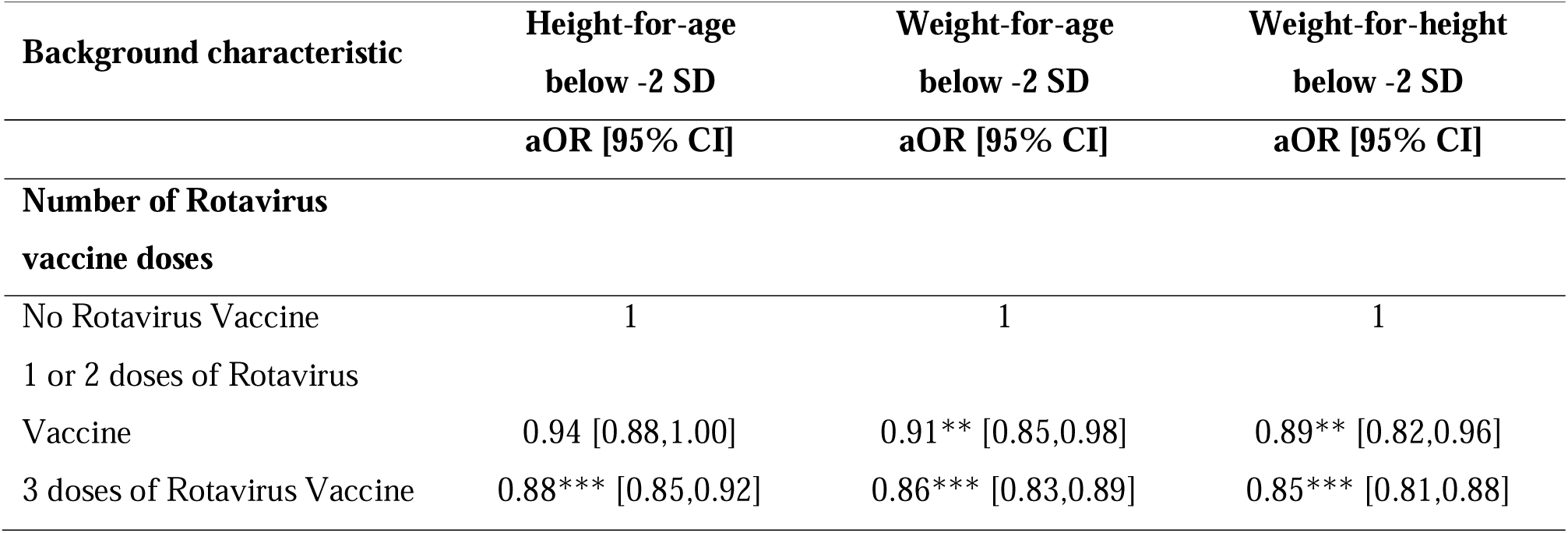
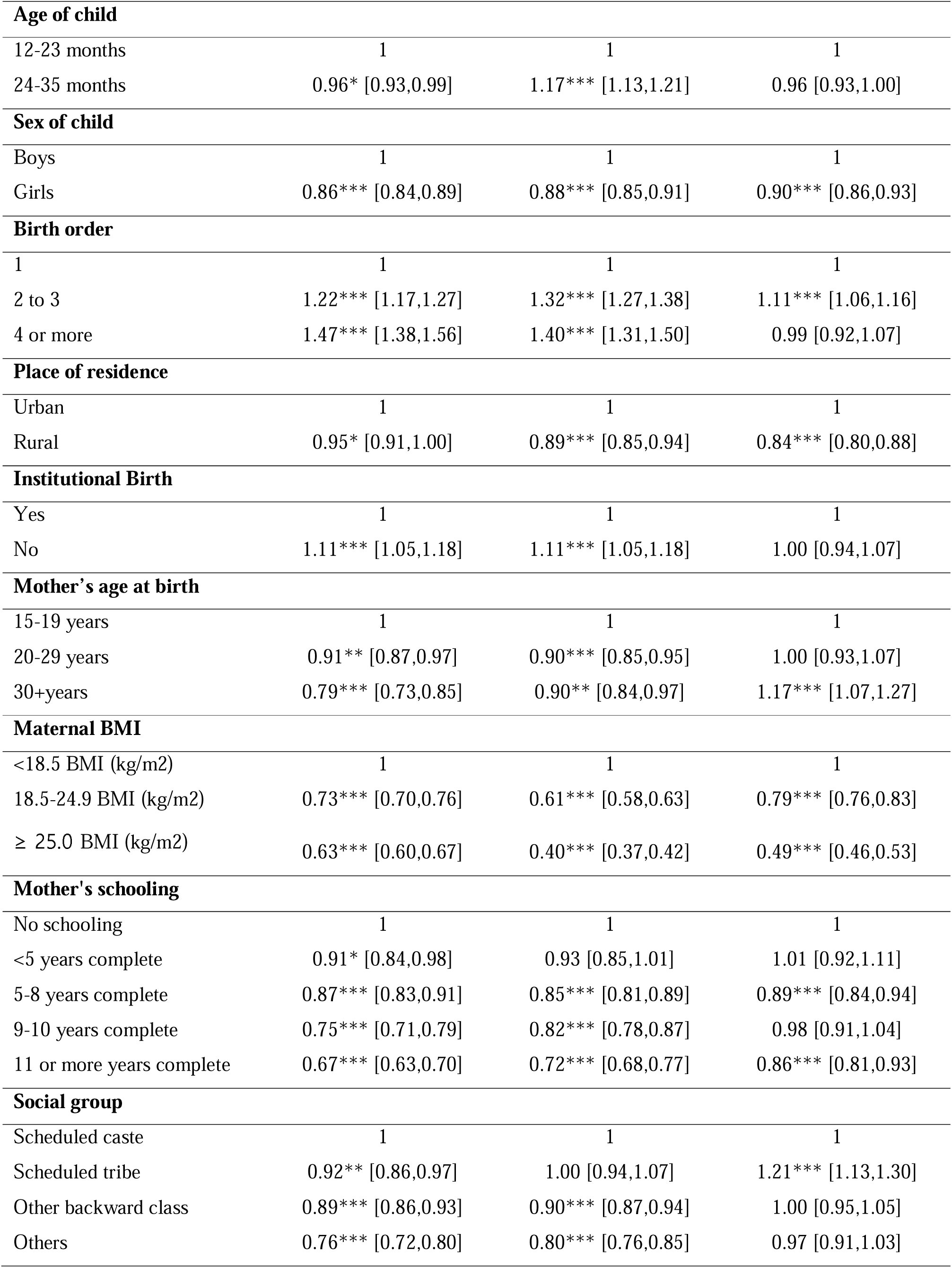
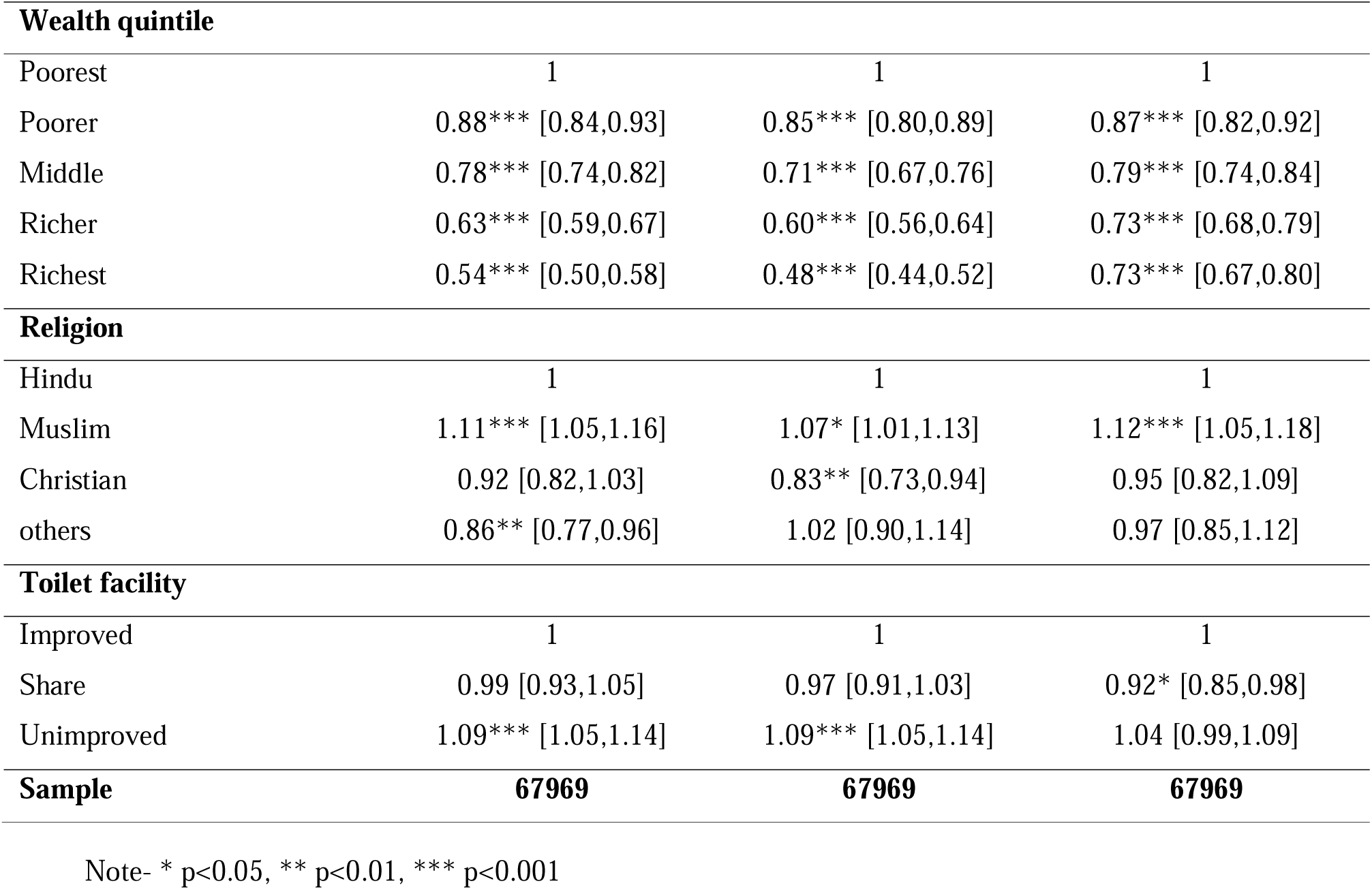
Multivariate logistic regression of Rotavirus vaccine and Undernutrition among children below –2 SD by sociodemographic characteristics in India, NFHS-5 (2019-21)

### 3.4 Regression analysis of Rotavirus vaccination and interaction of diarrhea & undernutrition

Table 3 shows the relationships between rotavirus vaccination and the concurrent incidence of diarrhea and undernutrition in children. This finding shows how no vaccination, partial vaccination (1 or 2 doses), and complete vaccination (all three doses) are differently associated with stunting (height-for-age), underweight (weight-for-age), and wasting (weight-for-height) in combination with diarrhea. Children who received three doses of the RVV showed significantly lower odds of experiencing both diarrhea and stunting (aOR = 0.74, 95% CI: 0.67–0.82), diarrhea and underweight (aOR = 0.71, 95% CI: 0.64–0.79), and diarrhea and wasting (aOR = 0.63, 95% CI: 0.55–0.73) compared to those who were unvaccinated.

**Table 3:** Association between Rotavirus vaccines, Diarrhea & Undernutrition among children.

| **Background characteristic** | **Diarrhea & Height-for-age below -2 SD** | **Diarrhea & Weight-for-age below -2 SD** | **Diarrhea & Weight-for-height below -2 SD** |
| --- | --- | --- | --- |
|  | **aOR [95% CI]** | **aOR [95% CI]** | **aOR [95% CI]** |
| **Number of Rotavirus vaccine doses** |  |  |  |
| No Rotavirus Vaccine | 1 | 1 | 1 |
| 1 or 2 doses of Rotavirus Vaccine | 0.77** [0.64,0.92] | 0.77* [0.63,0.94] | 0.62*** [0.47,0.82] |
| 3 doses of Rotavirus Vaccine | 0.74*** [0.67,0.82] | 0.71*** [0.64,0.79] | 0.63*** [0.55,0.73] |

Note- \* p<0.05, \*\* p<0.01, \*\*\* p<0.001; Adjusted variables are age of child, sex of child, birth order, place of residence, institutional birth, mother's age at birth, maternal BMI, mother's schooling, social group, wealth quintile, religion, and toilet facility.

### 3.5 Sensitivity analysis of the indirect effect of Rotavirus vaccination on child nutrition

We assessed the indirect effects of rotavirus vaccination for one or two doses or as a full three-dose schedule on stunting, underweight, and wasting among children aged 12-35 months. The indirect effects represent the RVV’s impact on children’s nutritional status via the mediation pathway through diarrhea reduction (Table 4). Among children who received one or two doses of the RVV, as well as those who received three doses, the indirect effects on the low risk of stunting (height-for-age Z-score < –2 SD), underweight (weight-for-age Z-score < –2 SD), and wasting (weight-for-height Z-score < –2 SD) through reduced incidence of diarrhea were not statistically significant. However, the indirect effect of the full three-dose RVV, via reduction in diarrhea, was significantly associated with a lower risk of underweight (OR = 0.98; 95% CI: 0.97–0.99) and wasting (OR = 0.98; 95% CI: 0.97–1.00), while not significant for stunting.

**Table 4:**
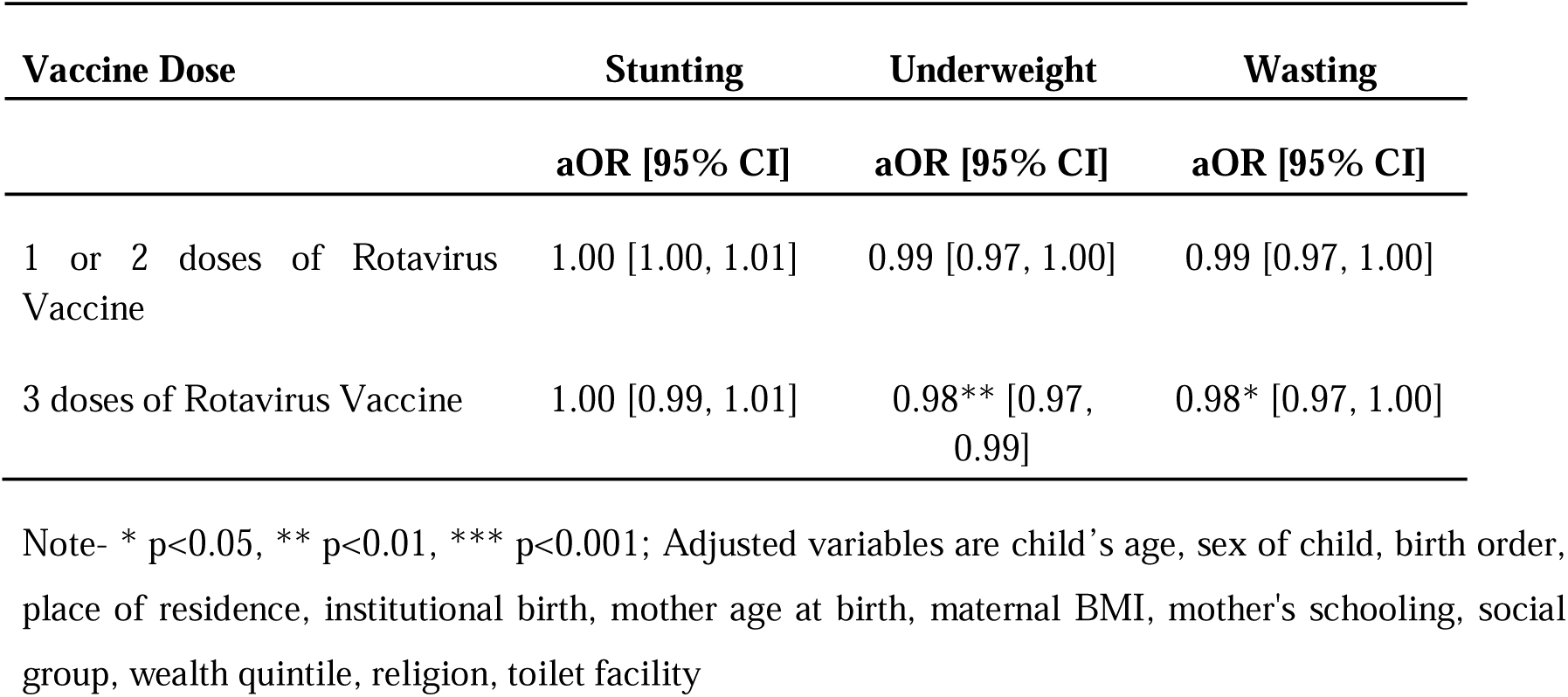
Mediation effect of the rotavirus vaccine via reduced diarrhea on undernutrition outcomes among children aged 12-35 months, India.

| Vaccine Dose | Stunting | Underweight | Wasting |
| --- | --- | --- | --- |
|  | aOR [95% CI] | aOR [95% CI] | aOR [95% CI] |
| 1 or 2 doses of Rotavirus Vaccine | 1.00 [1.00, 1.01] | 0.99 [0.97, 1.00] | 0.99 [0.97, 1.00] |
| 3 doses of Rotavirus Vaccine | 1.00 [0.99, 1.01] | 0.98** [0.97, 0.99] | 0.98* [0.97, 1.00] |

Note- \* p<0.05, \*\* p<0.01, \*\*\* p<0.001; Adjusted variables are child's age, sex of child, birth order, place of residence, institutional birth, mother age at birth, maternal BMI, mother's schooling, social group, wealth quintile, religion, toilet facility

### 3.6 Inverse probability weighted regression adjustment

Table 5 presents inverse probability weighted regression adjustment (IPWRA) estimates of the average treatment effect (ATE) and the average treatment effect on the treated (ATT) for rotavirus vaccination and child malnutrition outcomes among children aged 12-35 months in India. Children who received any doses of the rotavirus vaccine were linked to a 2.6%-point reduction in stunting (ATT= –0.026, 95% CI: –0.034 to –0.018), a 2.8%-point reduction in underweight (ATT= –0.028, 95% CI: –0.035 to –0.02), and a 2.3%-point reduction in wasting (ATT= –0.023, 95% CI: –0.03 to –0.017). The results of model 2 were also similar when restricting the treatment group to children who received all three doses of the vaccine. Figure 4 shows the kernel density of the propensity score distribution for covariates by rotavirus vaccine status in both models I and II. It also displays the distribution of a covariate propensity score after matching (ATT) was balanced compared to before matching.

**Figure 4:**
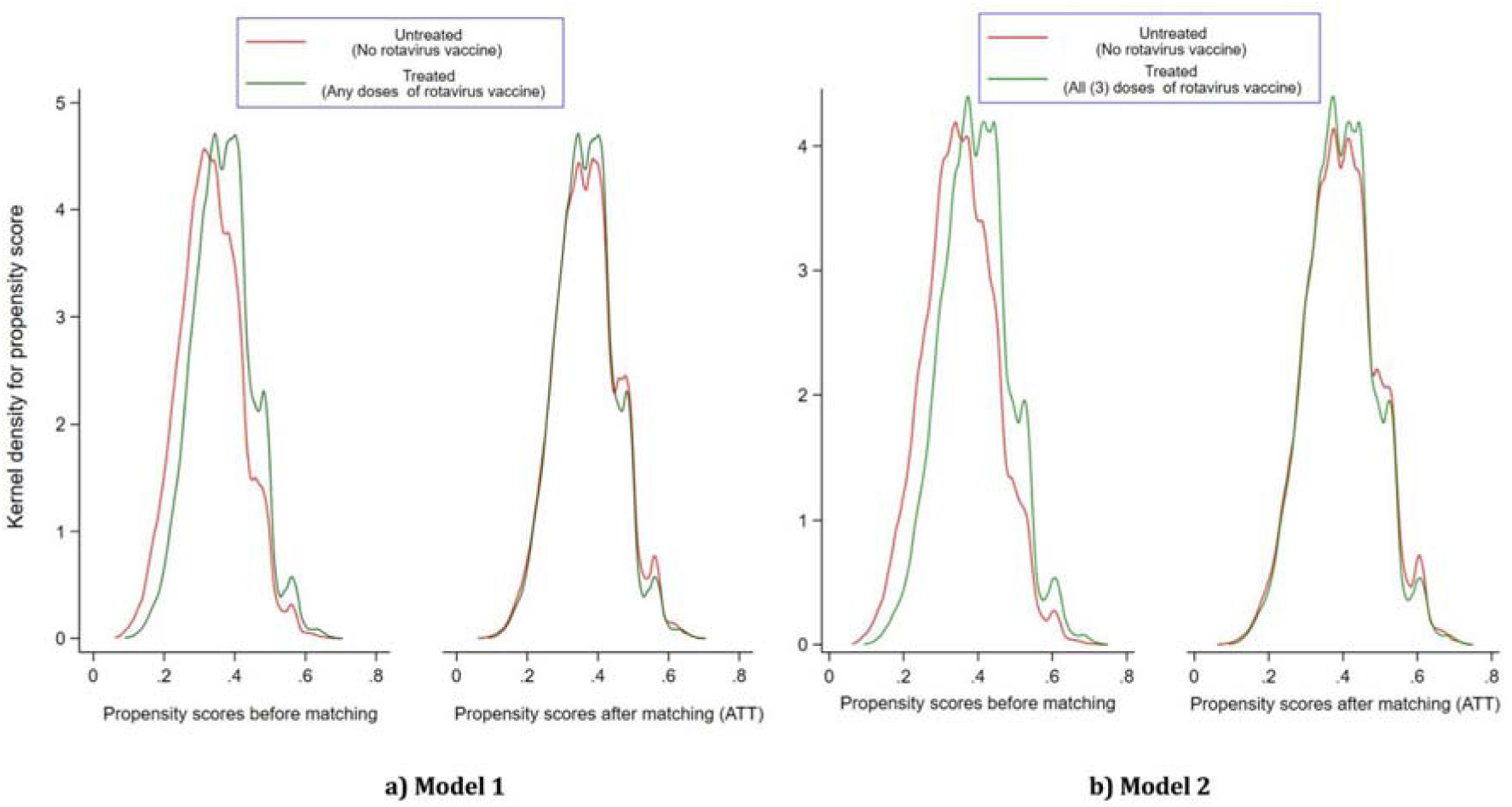
Propensity score distribution by zero and any doses of rotavirus vaccine.

**Table-5:**
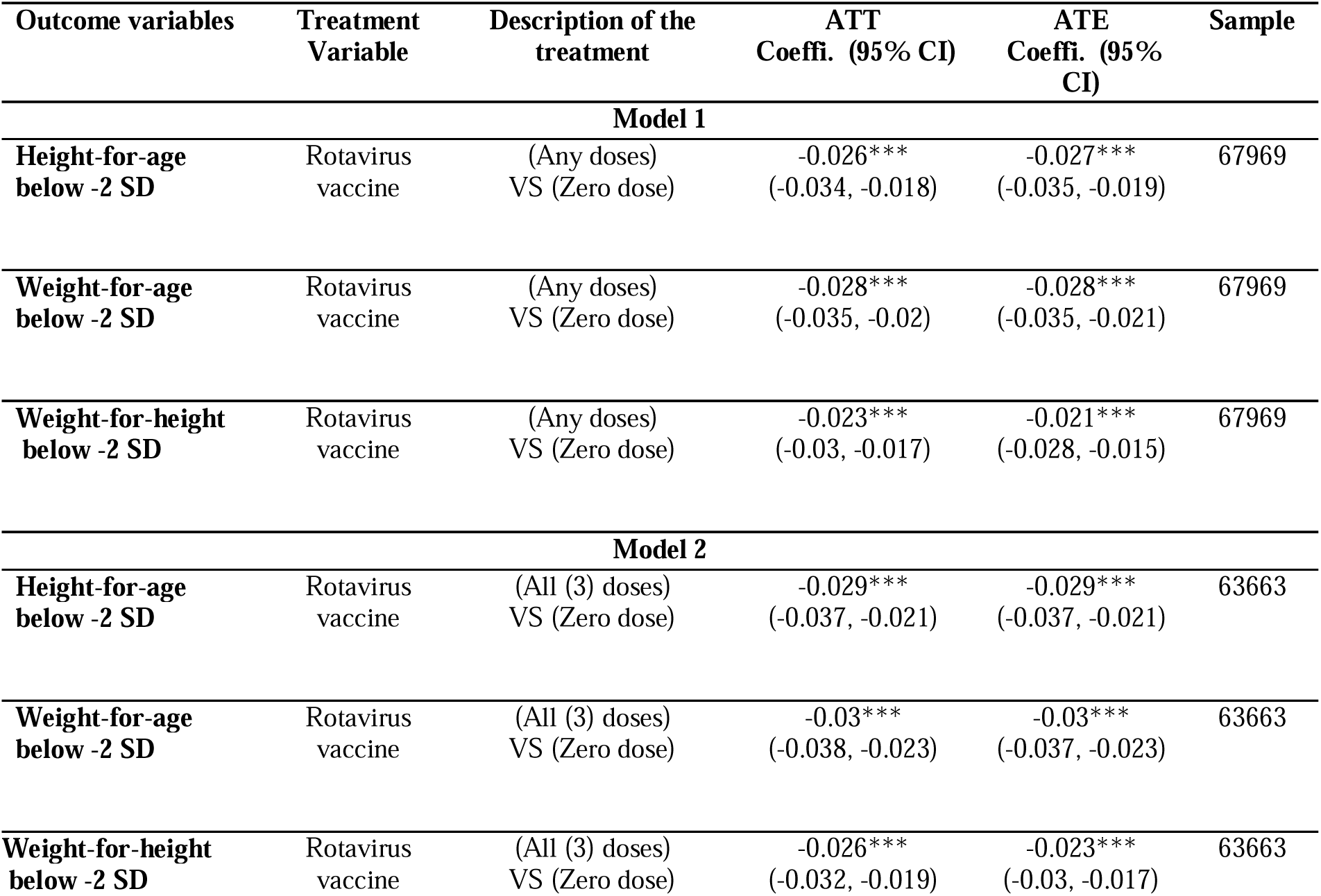
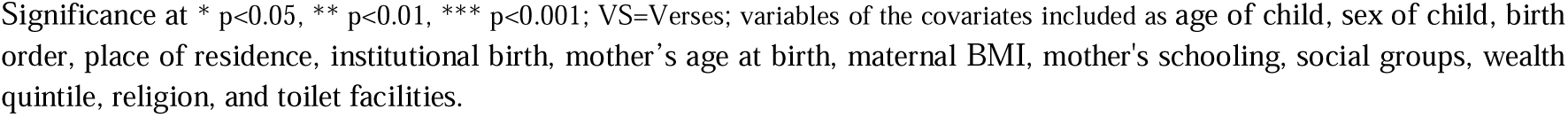
Inverse-probability-weighted regression adjustment (IPWRA) estimates of average treatment effect and average treatment effect on the treated.

## 4. Discussion

This study investigated the link between the RVV and malnutrition among children aged 12-35 months in India. The RVV is highly effective in addressing malnutrition in children under 5. The analysis revealed five key findings related to stunting, underweight, and wasting among children aged 12-35 months. First, receiving three doses of the RVV significantly reduced the likelihood of moderate and severe malnutrition across all three indicators: stunting, underweight, and wasting. Second, vaccinated children also had a lower risk of concurrent diarrheal infection and undernutrition. Third, certain sociodemographic variables had a significant impact on children’s malnutrition status. Children from rural areas had a lower risk of malnutrition than those in urban areas. Factors such as maternal education, mother’s age during childbirth, maternal BMI, and household wealth status had a significant impact on child malnutrition. The fourth finding emerged from the sensitivity analysis, a two-step statistical method to understand how the RVV may reduce undernutrition in children by preventing diarrhea. We first examined the direct link between vaccination and reduced diarrhea, then assessed how this reduction in diarrhea helped improve children’s nutritional status. This approach allows us to measure the vaccine’s indirect benefits and how it enhances growth by reducing illnesses that interfere with nutrient absorption. The findings highlight that, although the vaccine does not directly improve nutrition, it plays a critical role by breaking the cycle of frequent diarrhea and poor growth in children. The fifth finding is the treatment effect of RVV on malnutrition, showing that receiving any dose of RVV reduces the risk of stunting, underweight, and wasting among children aged 12-35 months.

Children who received all three doses of the RVV within their first year of life had notably better nutritional outcomes. Specifically, their risk of being moderately stunted, underweight, or wasted (below –2 SD) was reduced by 12%, 14%, and 15%, respectively. The protective effect of the vaccine was also evident in more severe cases of malnutrition, with significant reductions in the likelihood of falling below –3 SD across all three indicators. This highlights the importance of completing the recommended vaccination schedule, as children who are not fully vaccinated may be more vulnerable to diarrheal diseases and associated nutritional challenges. Interestingly, the analysis revealed that children in rural areas were less likely to experience moderate or severe growth failure than those in urban areas. These results align with other studies that found that children under age five from rural India had a lower probability of stunting, underweight, and wasting[46–48] [46–48]. Rural mothers typically demonstrate higher rates of early initiation and exclusive breastfeeding, as well as longer breastfeeding durations. They also have better access to Integrated Child Development Services (ICDS), dietary diversity, and seasonal, nutritious foods than urban mothers[46,47,49]. Urban mothers, particularly those living in urban slums and who frequently migrate, have challenging times in fulfilling their children’s dietary requirements[50].

Additionally, we found that higher levels of maternal education, higher wealth status, and mothers with normal BMI are associated with a lower likelihood of children being moderately or severely stunted, underweight, or wasted. This study’s findings are consistent with multiple studies that have established that mothers with higher education levels, higher household income, and better maternal nutrition have significantly lower odds of having stunted or underweight children compared to those with little or no education[46,51–53]. This stems from the significant role of mothers as primary caregivers and decision-makers for their children’s nutrition, apart from vaccination and overall development.

The effects of rotavirus vaccination on malnutrition in children have been mixed in previous research. However, our findings are consistent with a study conducted in Peru[31], which found that after adjusting for demographic, health, and household characteristics, children under five years who received the RVV had a mean height-for-age z-score (HAZ) that was 0.06 standard deviations higher than those who did not receive the vaccine. In contrast, a clinical trial conducted in Bangladesh did not show a significant effect of rotavirus vaccination on malnutrition indicators in children aged 27 to 38 months[54]. This lack of effect may be due to the vaccine’s low efficacy against severe rotavirus gastroenteritis in that population. A pooled analysis of studies from five LMICs demonstrated the cumulative effects of repeated diarrheal episodes from 0 to 24 months. For every five episodes of diarrhea that a child experiences, they are 13% more likely to be stunted at age two[28]. Consequently, the vaccine may not sufficiently reduce the overall burden of diarrheal illnesses among all recipients. Studies suggest that the effectiveness of rotavirus vaccines, which protect against a common cause of childhood diarrhea, may be influenced by children’s nutritional status. Clinical research has shown the significant efficacy of the rotavirus vaccine in reducing the risk of rotavirus disease in both well-nourished and malnourished children[55–57].

It has been widely accepted and generally considered a democratic pathogen because every child under age five is infected by the rotavirus[58]. Once infected, there is a substantial risk of developing mild to severe episodes of diarrhea, leading to malabsorption and malnutrition. In India, rotavirus is estimated to account for nearly 40% of moderate-to-severe diarrhea cases in children under age five. Furthermore, secondary data analysis indicates that children aged 12 to 35 months who received all three doses of RVV are significantly less likely to experience diarrheal disease, and there is a significant decline in overall diarrheal prevalence after introduction of the RVV under UIP[17]. Thus, the RVV plays a crucial role in preventing diarrheal diseases in children, ultimately supporting their physical growth as measured by height-for-age, weight-for-age, and weight-for-height. Since malnutrition is both a major risk factor for and a consequence of diarrheal disease (bidirectional), improving nutritional status by preventing diarrheal disease through the RVV is essential. The RVV breaks the cycle of diarrhea and malnutrition by directly preventing rotavirus infection and indirectly protecting against other pathogens causing diarrhea, a major cause of moderate to severe diarrhea in children. This reduces the burden of diarrheal illness, thereby decreasing malnutrition by improving gut malabsorption and appetite. By preventing rotavirus-related diarrhea in children, the vaccine improves nutrient absorption and maintains nutritional status, effectively breaking the vicious cycle. Rotavirus infections significantly impair nutritional status by disrupting both nutrient intake and absorption, increasing catabolism, and altering the physiological use of nutrients essential for growth and tissue repair.

Diarrheal diseases, in particular, contribute to malnutrition through multiple pathways. This impaired gut function contributes to a vicious cycle in which poor nutritional status increases susceptibility to infections, and repeated infections further aggravate malnutrition. The reduced dietary intake due to illness-related anorexia damages the intestinal mucosa, thereby decreasing the absorptive surface, and causes direct loss of nutrients through gastrointestinal secretions[59]. These effects are often accompanied by reduced gut enzymatic activity and a heightened catabolic response that further depletes micronutrient and macronutrient stores. Among the various pathogens responsible for diarrhea, rotavirus is consistently identified as the leading cause of severe diarrheal episodes in CU5[22]. Unlike some other enteric pathogens, rotavirus has a more pronounced impact on gut integrity, leading to villous atrophy and dysbiosis that impair nutrient absorption, particularly proteins and calories. This contributes directly to growth faltering and undernutrition. Evidence from longitudinal field studies, including those conducted by Leonardo Mata and colleagues at the Institute of Nutrition of Central America and Panama, has illustrated the cyclical nature of infection and malnutrition[60,61]. Their findings documented how repeated episodes of diarrhea closely align with periods of linear growth faltering in children. Given this link, rotavirus vaccination plays a crucial indirect role in addressing childhood malnutrition. By preventing or mitigating the severity of rotavirus-induced diarrhea, the vaccine reduces the frequency and duration of enteric insults to the gut, preserving absorptive function and supporting better nutrient utilization. This indirect benefit enhances children’s overall nutritional status and contributes to improved anthropometric outcomes over time. The introduction of the RVV represents a critical public health intervention capable of interrupting this cycle. Thus, beyond its primary role in reducing diarrheal morbidity and mortality, rotavirus vaccination is essential for breaking the vicious cycle between infection and malnutrition.

The RVV has demonstrated robust heterologous effects, providing benefits beyond protection against rotavirus alone. By significantly reducing the overall incidence of diarrheal disease, including cases caused by non-rotavirus pathogens, it serves as a critical intervention in low-resource settings where enteric infections and undernutrition frequently overlap[62]. Fewer diarrheal episodes translate into reduced gut inflammation, improved nutrient absorption, and a meaningful break in the cycle linking repeated infections to childhood malnutrition. This broader impact is primarily attributed to the vaccine’s ability to enhance mucosal immunity. Specifically, it stimulates gut-associated lymphoid tissue (GALT), triggering the development of memory B and T cells that can rapidly respond to subsequent infections, even when caused by unrelated pathogens. Furthermore, the vaccine promotes the production of immunoglobulin A (IgA) in the intestinal lining, a key antibody that helps neutralize pathogens and maintain gut barrier function[63]. Through this mucosal priming, the RVV not only protects against severe rotavirus diarrhea but also fortifies the gut’s immune defense against a wide range of enteric pathogens[64–67].

Addressing malnutrition requires a comprehensive approach integrating nutritional interventions with other community-based programs. Like other vaccinations, the RVV is cost-effective and provides a positive return on investment for both governments and families. Emerging evidence increasingly supports that the impact of rotavirus infection extends beyond gastrointestinal illness, with significant implications for child growth and nutritional outcomes. In a few recent studies, it has been found that a considerable proportion of rotavirus cases were observed among children with moderate to severe malnutrition. Conversely, malnourished children are more susceptible to repeated and severe diarrheal episodes, perpetuating a chain, reinforcing the bi-directional relationship between diarrheal disease and undernutrition. Given the strong evidence linking rotavirus vaccination to better nutritional outcomes, it is essential that pediatricians and medical practitioners across the country actively promote and ensure that all eligible children receive all three recommended doses of the vaccine. In India, immunization and nutrition fall under the purview of the Ministry of Health & Family Welfare (MoHFW) and the Ministry of Women and Child Development (MoWCD), respectively. National Nutrition Mission, launched in 2018 under MoWCD, was an integration effort between the two ministries, emphasizing nutrition and immunization as key strategies to reduce child malnutrition. Anganwadi workers (AWW), the frontline health workers in India—the first and most frequent point of contact for families with young children, conduct frequent awareness drives on the benefits of the RVV in communities. This is essential for improving RVV coverage. Since the AWW already delivers key services related to nutrition, growth monitoring, and Water, Sanitation, and Hygiene (WASH), their engagement in vaccine promotion could make a real difference. Improved coordination with the health department and frontline workers can help ensure children complete their RVV schedule alongside routine care. At the same time, Urban Local Bodies (ULB), the key implementers of the National Urban Health Mission (NUHM, a sub-mission of the National Health Mission), have a significant role in providing primary health services. Under the NUHM, ULBs are responsible for tackling diarrhea and malnutrition in cities, two challenges that the RVV helps to address. They must be supported and held accountable to maintain strong vaccine coverage. The recent NFHS data have shown relatively higher diarrheal prevalence in urban regions, and the findings of this study also call for greater attention in urban areas. It is recommended that the RVV be included as a process indicator in health and nutrition interventions led by ULBs. Rotavirus vaccine uptake can be a meaningful measure of progress, reducing out-of-pocket costs amongst the urban poor.

By preventing frequent and severe diarrheal episodes, the vaccine not only reduces under-five mortality quantitatively but also qualitatively improves child health in LMICs by breaking the cycle of infection-induced malabsorption and growth faltering. This dual effect positions the RVV as a powerful intervention that enhances both survival and healthy development in early childhood. It can be concluded that rotavirus vaccination reduces diarrhea-associated mortality and improves gut health and nutritional outcomes in young children. Thus, the RVV contributes not only to reducing diarrheal morbidity but also to improved nutritional outcomes and healthier growth trajectories in children.

## 5. Limitations

Along with important findings, this study has a few limitations. First, the analysis relies on cross-sectional household survey data, which inherently limits the ability to establish temporal relationships as compared to clinical treatments and laboratory experiments, which involve controlled, repeated measurements over time. As a result, we were unable to identify specific enteric pathogens associated with diarrheal episodes among acutely malnourished children. The lack of clinical data prevents us from distinguishing between types of diarrheal infections or assessing their severity. Second, our analysis does not account for potential confounding factors such as the use of Oral Rehydration Solutions (ORS), zinc supplementation, or timely access to healthcare services, all of which could influence both diarrheal incidence and nutritional outcomes. The absence of information on rotavirus-specific infection or diarrheal energy loss further limits our understanding of the precise biological mechanisms linking rotavirus vaccination and improved growth indicators. Lastly, while the study provides suggestive associations between rotavirus vaccination and reduced malnutrition, more robust longitudinal evidence is needed to validate these findings and quantify the directionality of the effects. Future research incorporating prospective cohorts or randomized controlled trials can be conducted to explore potential causal inference and adjust for unmeasured confounders.

## 6. Conclusion

This study offers important novel evidence from India on the association between rotavirus vaccination and key indicators of child malnutrition. In a context where malnutrition remains a persistent public health challenge, particularly among children under five, our findings suggest that the RVV may serve as a valuable intervention to break the well-documented vicious cycle between diarrhea and undernutrition. By preventing rotavirus-induced diarrheal episodes directly and indirectly reducing the burden of other diarrheal pathogens, the vaccine contributes not only to reducing child morbidity and mortality but also to supporting improved nutritional outcomes. This is especially relevant for low– and middle-income countries like India, where diarrheal diseases continue to exert a heavy burden on the public health system. The findings also align with the broader objectives of the Government of India’s flagship POSHAN Abhiyaan, which addresses malnutrition through a multi-sectoral approach. Expanding RVV coverage nationwide represents a key strategy under this mission and has the potential to produce measurable improvements in child health and nutrition outcomes.

## Authors Contributors

Conceptualisation: Pritu Dhalaria, Ajay Kumar Verma, and Pawan Kumar. Methodology: Ajay Kumar Verma and Ajeet Kumar Singh. Formal analysis: Ajay Kumar Verma and Ajeet Kumar Singh. Investigation: Pritu Dhalaria, Kapil Singh, and Pretty Priyadarshini. Writing—original draft preparation: Ajay Kumar Verma, Pretty Priyadarshini, and Ajeet Kumar Singh. Writing—review and editing: Pretty Priyadarshini, Ajeet Kumar Singh, Pawan Kumar, and Bhupendra Tripathi. Visualisation: Pretty Priyadarshini and Ajeet Kumar Singh. Supervision: Pritu Dhalaria, Pawan Kumar, Sanjay Kapur, Arindam Ray, and Bhupendra Tripathi. Guarantor information: Ajay Kumar Verma accepts full responsibility for the finished work and/or the conduct of the study, has access to the data, and controls the decision to publish.

## Data availability statement

Data are available in a public, open-access repository. The data are available in the public domain and can be downloaded on reasonable request. https://dhsprogram.com/data/available-datasets.cfm

## Ethics Statement

The study is based on secondary household survey data available in the public domain, with no primary data collection and no human participants involved directly in it; hence, no ethical approval is required.

