## Supplementary Tables for "Impact of Rotavirus Vaccine on Malnutrition Among Children in India—Breaking Vicious Cycle of Diarrhea: A Cross-Sectional Analytical Study"

**Supplementary Table -1: Multivariate logistic regression of Rotavirus vaccine and Undernutrition among children below -3 SD by sociodemographic characteristics in India, NFHS-5 (2019-21)**

| **Background characteristic** | **Height-for-age**  **below -3 SD** | **Weight-for-age**  **below -3 SD** | **Weight-for-height below -3 SD** |
| --- | --- | --- | --- |
|  | **aOR [95% CI]** | **aOR [95% CI]** | **aOR [95% CI]** |
| **Number of Rotavirus vaccine doses** |  |  |  |
| No Rotavirus Vaccine | 1 | 1 | 1 |
| 1 or 2 doses of Rotavirus Vaccine | 0.99 [0.91,1.07] | 1.02 [0.92,1.14] | 0.95 [0.85,1.07] |
| 3 doses of Rotavirus Vaccine | 0.86*** [0.82,0.90] | 0.85*** [0.80,0.90] | 0.82*** [0.77,0.87] |
| **Age of child** |  |  |  |
| 12-23 months | 1 | 1 | 1 |
| 24-35 months | 0.82*** [0.79,0.86] | 1.05 [0.99,1.11] | 0.97 [0.91,1.03] |
| **Sex of child** |  |  |  |
| Boys | 1 | 1 | 1 |
| Girls | 0.83*** [0.80,0.86] | 0.91*** [0.86,0.96] | 0.90*** [0.85,0.96] |
| **Birth order** |  |  |  |
| 1 | 1 | 1 | 1 |
| 2 to 3 | 1.15*** [1.09,1.21] | 1.32*** [1.23,1.41] | 1.08* [1.01,1.16] |
| 4 or more | 1.40*** [1.30,1.51] | 1.48*** [1.34,1.63] | 0.96 [0.86,1.08] |
| **Place of residence** |  |  |  |
| Urban | 1 | 1 | 1 |
| Rural | 0.85*** [0.81,0.90] | 0.85*** [0.78,0.91] | 0.84*** [0.78,0.90] |
| **Institutional birth** |  |  |  |
| Yes | 1 | 1 | 1 |
| No | 1.17*** [1.10,1.25] | 1.23*** [1.13,1.33] | 1.01 [0.92,1.11] |
| **Mother's age at birth** |  |  |  |
| 15-19 years | 1 | 1 | 1 |
| 20-29 years | 0.92* [0.86,0.98] | 0.88** [0.81,0.97] | 1.01 [0.92,1.12] |
| 30+years | 0.79*** [0.72,0.87] | 0.89 [0.79,1.00] | 1.32*** [1.17,1.50] |
| **Maternal BMI** |  |  |  |
| <18.5 BMI (kg/m2) | 1 | 1 | 1 |
| 18.5-24.9 BMI (kg/m2) | 0.80*** [0.77,0.84] | 0.65*** [0.61,0.69] | 0.92* [0.86,0.99] |
| ≥ 25.0 BMI (kg/m2) | 0.65*** [0.60,0.69] | 0.41*** [0.37,0.45] | 0.55*** [0.50,0.61] |
| **Mother's schooling** |  |  |  |
| No schooling | 1 | 1 | 1 |
| <5 years complete | 0.84*** [0.77,0.93] | 0.93 [0.83,1.05] | 0.94 [0.81,1.08] |
| 5-8 years complete | 0.81*** [0.76,0.86] | 0.77*** [0.71,0.83] | 0.85*** [0.78,0.93] |
| 9-10 years complete | 0.66*** [0.62,0.71] | 0.74*** [0.68,0.81] | 0.87** [0.79,0.96] |
| 11 or more years complete | 0.57*** [0.53,0.61] | 0.63*** [0.57,0.69] | 0.85** [0.77,0.94] |
| **Social group** |  |  |  |
| Scheduled caste | 1 | 1 | 1 |
| Scheduled tribe | 0.98 [0.91,1.05] | 1.13** [1.04,1.24] | 1.23*** [1.11,1.37] |
| Other backward class | 0.92** [0.87,0.97] | 0.90** [0.85,0.97] | 1.00 [0.93,1.08] |
| Others | 0.86*** [0.81,0.92] | 0.79*** [0.72,0.86] | 1.06 [0.97,1.16] |
| **Wealth quintile** |  |  |  |
| Poorest | 1 | 1 | 1 |
| Poorer | 0.85*** [0.80,0.90] | 0.82*** [0.76,0.88] | 0.96 [0.88,1.05] |
| Middle | 0.74*** [0.69,0.79] | 0.68*** [0.63,0.75] | 0.89* [0.80,0.98] |
| Richer | 0.58*** [0.53,0.62] | 0.55*** [0.49,0.61] | 0.87* [0.78,0.98] |
| Richest | 0.53*** [0.48,0.58] | 0.47*** [0.41,0.54] | 0.86* [0.75,0.98] |
| **Religion** |  |  |  |
| Hindu | 1 | 1 | 1 |
| Muslim | 1.12*** [1.05,1.19] | 1.11* [1.02,1.20] | 1.11* [1.02,1.21] |
| Christian | 0.95 [0.82,1.11] | 0.62*** [0.49,0.78] | 0.83 [0.67,1.03] |
| Others | 0.95 [0.82,1.10] | 0.93 [0.76,1.14] | 0.95 [0.77,1.16] |
| **Toilet facility** |  |  |  |
| Improved | 1 | 1 | 1 |
| Share | 1.02 [0.94,1.09] | 0.96 [0.87,1.06] | 0.96 [0.86,1.07] |
| Unimproved | 1.03 [0.98,1.09] | 1.10** [1.03,1.17] | 1.04 [0.97,1.12] |
| **Sample** | **67969** | **67969** | **67969** |

Note- * p<0.05, ** p<0.01, *** p<0.001

Table-1 presents the results of the association between rotavirus vaccination and severe malnutrition outcomes. Severe malnutrition refers to height-for-age, weight-for-age, and weight-for-height measurements below -3 standard deviations (SD) below the mean. In contrast, children who received all three doses of rotavirus vaccines demonstrated a significant reduction in the odds of severe malnutrition across all three outcomes: height-for-age (aOR: 0.86, 95% CI: 0.82–0.90), weight-for-age (aOR: 0.85, 95% CI: 0.80–0.90), and weight-for-height (aOR: 0.82, 95% CI: 0.77–0.87). Girl child, children from rural areas, children of mothers with normal BMI, children of mothers who completed more than 10 years of schooling, and children from the highest wealth quintile had relatively lower odds of severe malnutrition outcomes. In contrast, children of higher birth order, history of non-institutional delivery, and those with unimproved toilet facilities had relatively higher odds of severe malnutrition outcomes.

**Supplementary Table-2: Multivariate logistic regression of Rotavirus vaccine and Undernutrition among children below -2 SD by sociodemographic characteristics among children 12-23 months who received all routine immunizations in India, NFHS-5 (2019-21)**

| **Background characteristic** | **Height-for-age**  **below -2 SD** | **Weight-for-age**  **below -2 SD** | **Weight-for-height  below -2 SD** |
| --- | --- | --- | --- |
|  | **OR [95% CI]** | **OR [95% CI]** | **OR [95% CI]** |
| **Number of Rotavirus vaccine doses** |  |  |  |
| No Rotavirus Vaccine | 1.00 [1.00,1.00] | 1.00 [1.00,1.00] | 1.00 [1.00,1.00] |
| 1 or 2 doses of Rotavirus Vaccine | 0.85* [0.74,0.97] | 0.94 [0.81,1.09] | 0.90 [0.76,1.06] |
| 3 doses of Rotavirus Vaccine | 0.90*** [0.85,0.95] | 0.87*** [0.82,0.92] | 0.82*** [0.77,0.88] |
| **Age of child** |  |  |  |
| 12-23 months | 1.00 [1.00,1.00] | 1.00 [1.00,1.00] | 1.00 [1.00,1.00] |
| 24-35 months | 1.33 [0.85,2.09] | 0.79 [0.47,1.33] | 0.85 [0.47,1.52] |
| **Sex of child** |  |  |  |
| Boys | 1.00 [1.00,1.00] | 1.00 [1.00,1.00] | 1.00 [1.00,1.00] |
| Girls | 0.81*** [0.77,0.85] | 0.79*** [0.75,0.84] | 0.92* [0.87,0.98] |
| **Birth order** |  |  |  |
| 1 | 1.00 [1.00,1.00] | 1.00 [1.00,1.00] | 1.00 [1.00,1.00] |
| 2 to 3 | 1.19*** [1.12,1.26] | 1.37*** [1.28,1.46] | 1.10* [1.02,1.18] |
| 4 or more | 1.40*** [1.26,1.55] | 1.37*** [1.22,1.53] | 1.02 [0.90,1.16] |
| **Place of residence** |  |  |  |
| Urban | 1.00 [1.00,1.00] | 1.00 [1.00,1.00] | 1.00 [1.00,1.00] |
| Rural | 0.95 [0.89,1.02] | 0.90** [0.83,0.97] | 0.88** [0.81,0.96] |
| **Institutional birth** |  |  |  |
| Yes | 1.00 [1.00,1.00] | 1.00 [1.00,1.00] | 1.00 [1.00,1.00] |
| No | 1.05 [0.95,1.16] | 1.21*** [1.10,1.34] | 1.15* [1.03,1.29] |
| **Mother's age at birth** |  |  |  |
| 15-19 years | 1.00 [1.00,1.00] | 1.00 [1.00,1.00] | 1.00 [1.00,1.00] |
| 20-29 years | 0.90* [0.82,0.99] | 0.86** [0.78,0.95] | 1.03 [0.92,1.15] |
| 30+years | 0.75*** [0.66,0.84] | 0.88 [0.78,1.00] | 1.10 [0.95,1.27] |
| **Maternal BMI** |  |  |  |
| <18.5 BMI (kg/m2) | 1.00 [1.00,1.00] | 1.00 [1.00,1.00] | 1.00 [1.00,1.00] |
| 18.5-24.9 BMI (kg/m2) | 0.77*** [0.72,0.81] | 0.60*** [0.56,0.64] | 0.76*** [0.71,0.82] |
| ≥ 25.0 BMI (kg/m2) | 0.66*** [0.61,0.72] | 0.42*** [0.38,0.46] | 0.52*** [0.47,0.58] |
| **Mother's schooling** |  |  |  |
| No schooling | 1.00 [1.00,1.00] | 1.00 [1.00,1.00] | 1.00 [1.00,1.00] |
| <5 years complete | 0.98 [0.85,1.12] | 0.90 [0.78,1.03] | 0.86 [0.74,1.01] |
| 5-8 years complete | 0.91* [0.83,0.98] | 0.88** [0.81,0.96] | 0.83*** [0.75,0.91] |
| 9-10 years complete | 0.79*** [0.72,0.86] | 0.82*** [0.74,0.90] | 0.89* [0.80,0.99] |
| 11 or more years complete | 0.74*** [0.68,0.81] | 0.74*** [0.67,0.81] | 0.78*** [0.70,0.87] |
| **Social group** |  |  |  |
| Scheduled caste | 1.00 [1.00,1.00] | 1.00 [1.00,1.00] | 1.00 [1.00,1.00] |
| Scheduled tribe | 0.97 [0.88,1.07] | 1.01 [0.91,1.12] | 1.16** [1.04,1.30] |
| Other backward class | 0.87*** [0.81,0.93] | 0.90** [0.84,0.97] | 1.01 [0.93,1.10] |
| Others | 0.76*** [0.70,0.83] | 0.82*** [0.75,0.90] | 1.01 [0.91,1.12] |
| **Wealth quintile** |  |  |  |
| Poorest | 1.00 [1.00,1.00] | 1.00 [1.00,1.00] | 1.00 [1.00,1.00] |
| Poorer | 0.90** [0.83,0.97] | 0.81*** [0.75,0.88] | 0.83*** [0.75,0.91] |
| Middle | 0.76*** [0.70,0.83] | 0.65*** [0.59,0.71] | 0.76*** [0.68,0.84] |
| Richer | 0.66*** [0.60,0.73] | 0.54*** [0.48,0.60] | 0.68*** [0.60,0.76] |
| Richest | 0.58*** [0.52,0.66] | 0.42*** [0.37,0.48] | 0.61*** [0.53,0.71] |
| **Religion** |  |  |  |
| Hindu | 1.00 [1.00,1.00] | 1.00 [1.00,1.00] | 1.00 [1.00,1.00] |
| Muslim | 1.15*** [1.06,1.25] | 1.05 [0.96,1.15] | 0.99 [0.90,1.10] |
| Christian | 0.90 [0.75,1.08] | 0.87 [0.71,1.08] | 1.06 [0.85,1.32] |
| Others | 1.03 [0.87,1.22] | 1.17 [0.97,1.41] | 0.95 [0.76,1.17] |
| **Toilet facility** |  |  |  |
| Improved | 1.00 [1.00,1.00] | 1.00 [1.00,1.00] | 1.00 [1.00,1.00] |
| Share | 1.00 [0.91,1.10] | 0.97 [0.88,1.07] | 0.93 [0.83,1.04] |
| Unimproved | 1.13*** [1.06,1.21] | 1.13*** [1.06,1.21] | 1.07 [0.99,1.16] |
| **Sample** | **26125** | **26125** | **26125** |

* p<0.05, ** p<0.01, *** p<0.001

Table-2- Positive association between rotavirus vaccination and children with height-for-age, weight-for-age, and weight-for-height below -2 standard deviations (SD). Findings reveal that children who received all three rotavirus vaccines had a lower risk of stunting (aOR: 0.90, 95% CI: 0.85–0.95), underweight (aOR: 0.87, 95% CI: 0.82–0.92), and wasting (aOR: 0.82, 95% CI: 0.77–0.88) compared to those who did not receive any doses among children aged 12-23 months and who had all basic vaccinations.

**Supplementary Table-3: Multivariate logistic regression of Rotavirus vaccine and Undernutrition among children below -2 SD and whose vaccine was reported by card by sociodemographic characteristics in India, NFHS-5(2019-21)**

| **Background characteristic** | **Height-for-age**  **below -2 SD** | **Weight-for-age**  **below -2 SD** | **Weight-for-height  below -2 SD** |
| --- | --- | --- | --- |
|  | **OR [95% CI]** | **OR [95% CI]** | **OR [95% CI]** |
| **Rotavirus vaccine doses** |  |  |  |
| No | 1.00 [1.00,1.00] | 1.00 [1.00,1.00] | 1.00 [1.00,1.00] |
| Rotavirus 1&2 | 0.94* [0.88,1.00] | 0.91** [0.85,0.98] | 0.89** [0.82,0.96] |
| Rotavirus all | 0.89*** [0.85,0.92] | 0.85*** [0.82,0.88] | 0.83*** [0.79,0.87] |
| **Age of child** |  |  |  |
| 12-23 months | 1.00 [1.00,1.00] | 1.00 [1.00,1.00] | 1.00 [1.00,1.00] |
| 24-35 months | 0.97 [0.94,1.00] | 1.18*** [1.14,1.22] | 0.97 [0.93,1.01] |
| **Sex of child** |  |  |  |
| male | 1.00 [1.00,1.00] | 1.00 [1.00,1.00] | 1.00 [1.00,1.00] |
| female | 0.86*** [0.83,0.89] | 0.89*** [0.86,0.92] | 0.91*** [0.87,0.95] |
| **Birth order** |  |  |  |
| 1 | 1.00 [1.00,1.00] | 1.00 [1.00,1.00] | 1.00 [1.00,1.00] |
| 2 to 3 | 1.22*** [1.17,1.27] | 1.32*** [1.27,1.38] | 1.09*** [1.04,1.14] |
| 4 or more | 1.48*** [1.39,1.58] | 1.40*** [1.30,1.49] | 0.97 [0.90,1.05] |
| **Place of residence** |  |  |  |
| Urban | 1.00 [1.00,1.00] | 1.00 [1.00,1.00] | 1.00 [1.00,1.00] |
| Rural | 0.95* [0.91,0.99] | 0.89*** [0.85,0.93] | 0.84*** [0.79,0.88] |
| **Institutional Birt**h |  |  |  |
| No | 1.00 [1.00,1.00] | 1.00 [1.00,1.00] | 1.00 [1.00,1.00] |
| Yes | 1.12*** [1.06,1.19] | 1.11*** [1.05,1.18] | 1.00 [0.94,1.07] |
| **Mother’s age at birth** |  |  |  |
| 15-19 years | 1.00 [1.00,1.00] | 1.00 [1.00,1.00] | 1.00 [1.00,1.00] |
| 20-29 years | 0.92** [0.87,0.97] | 0.91** [0.86,0.97] | 1.00 [0.94,1.08] |
| 30+years | 0.78*** [0.73,0.84] | 0.92* [0.85,1.00] | 1.19*** [1.09,1.30] |
| **Maternal BMI** |  |  |  |
| Thin | 1.00 [1.00,1.00] | 1.00 [1.00,1.00] | 1.00 [1.00,1.00] |
| Normal | 0.73*** [0.70,0.76] | 0.61*** [0.58,0.63] | 0.79*** [0.75,0.82] |
| Obese | 0.63*** [0.60,0.67] | 0.39*** [0.37,0.42] | 0.49*** [0.46,0.53] |
| **Mother's schooling** |  |  |  |
| No schooling | 1.00 [1.00,1.00] | 1.00 [1.00,1.00] | 1.00 [1.00,1.00] |
| <5 years complete | 0.91* [0.84,0.99] | 0.94 [0.86,1.02] | 1.03 [0.93,1.13] |
| 5-8 years complete | 0.86*** [0.81,0.90] | 0.86*** [0.81,0.90] | 0.91** [0.85,0.97] |
| 9-10 years complete | 0.75*** [0.71,0.79] | 0.82*** [0.78,0.87] | 0.97 [0.91,1.04] |
| More than years complete | 0.67*** [0.63,0.71] | 0.72*** [0.68,0.77] | 0.88*** [0.82,0.94] |
| **Social group** |  |  |  |
| Scheduled caste | 1.00 [1.00,1.00] | 1.00 [1.00,1.00] | 1.00 [1.00,1.00] |
| Scheduled tribe | 0.92** [0.86,0.98] | 1.01 [0.95,1.08] | 1.21*** [1.12,1.30] |
| Other backward class | 0.90*** [0.86,0.94] | 0.90*** [0.86,0.94] | 0.98 [0.93,1.04] |
| Others | 0.76*** [0.72,0.80] | 0.79*** [0.75,0.84] | 0.97 [0.91,1.03] |
| **Wealth quintile** |  |  |  |
| Poorest | 1.00 [1.00,1.00] | 1.00 [1.00,1.00] | 1.00 [1.00,1.00] |
| Poorer | 0.90*** [0.86,0.95] | 0.86*** [0.82,0.91] | 0.85*** [0.80,0.91] |
| Middle | 0.78*** [0.74,0.83] | 0.72*** [0.68,0.76] | 0.78*** [0.73,0.84] |
| Richer | 0.63*** [0.59,0.67] | 0.60*** [0.56,0.65] | 0.74*** [0.69,0.80] |
| Richest | 0.55*** [0.51,0.60] | 0.48*** [0.44,0.52] | 0.72*** [0.66,0.79] |
| **Religion** |  |  |  |
| Hindu | 1.00 [1.00,1.00] | 1.00 [1.00,1.00] | 1.00 [1.00,1.00] |
| Muslim | 1.09** [1.03,1.14] | 1.05 [0.99,1.11] | 1.12*** [1.05,1.19] |
| Christian | 0.95 [0.84,1.07] | 0.86* [0.75,0.98] | 0.96 [0.83,1.11] |
| others | 0.87* [0.78,0.98] | 1.03 [0.91,1.17] | 0.97 [0.85,1.12] |
| **Toilet facility** |  |  |  |
| Improved | 1.00 [1.00,1.00] | 1.00 [1.00,1.00] | 1.00 [1.00,1.00] |
| Share | 0.98 [0.92,1.04] | 0.97 [0.91,1.03] | 0.91* [0.84,0.98] |
| Unimproved | 1.10*** [1.06,1.15] | 1.09*** [1.05,1.14] | 1.03 [0.98,1.08] |
| **Sample** | **63703** | **63703** | **63703** |

* p<0.05, ** p<0.01, *** p<0.001

Table 3 shows the results of a multivariate logistic regression examining the relationship between the Rotavirus vaccine and undernutrition below -2 SD using restricted data on vaccination, which was not reported among children aged 12-35 months by vaccination or health records. The results suggets that children who received all three doses of the rotavirus vaccine had a lower risk of stunting (aOR: 0.89, 95% CI: 0.85–0.92), underweight (aOR: 0.85, 95% CI: 0.82–0.98), and wasting (aOR: 0.83, 95% CI: 0.79–0.87) compared to children who did not receive any doses.
